# Endline Assessment of a Community-Based Program on Hypertension and Diabetes Management in Brazil

**DOI:** 10.1101/2022.05.22.22275385

**Authors:** Luisa Sorio Flor, Shelley Wilson, Welma Wildes Amorim, Mark TU Barone, Vanessa Moraes Bezerra, Paurvi Bhatt, Maria A Loguercio Bouskela, Joseph N Camarda, Christiane CR Cimini, Matheus L Cortes, Jessica Daly, Patrick W Endlich, Nancy Fullman, Katie Panhorst Harris, Clavdia N Kochergin, Marcia Maria Oliveira Lima, José A Louzado, Junia X Maia, Milena S Marcolino, Claire R McNellan, Danielle Souto de Medeiros, Sostenes Mistro, Marie Ng, Joao AQ Oliveira, Marcio Galvão Oliveira, Bryan K Phillips, Vânia S de O e Almeida Pinto, Antonio Luiz P Ribeiro, Davi Rumel, Kelle Oliveira Silva, Daniela Arruda Soares, Blake Thomson, Emmanuela Gakidou

## Abstract

**Background:** Brazil HealthRise community-based program focused on improving technologies for care coordination, developing the local workforce, and identifying and educating individuals with hypertension and diabetes.

**Objectives:** To assess the impact of HealthRise on hypertension and diabetes management among patients in the region of Teófilo Otoni (TO) and in the city of Vitória da Conquista (VC).

**Methods:** Grantees routinely collected patient-level clinical in intervention areas from March 2017 to December 2018; endline qualitative interviews were conducted with patients, providers, administrators, and policymakers in both intervention and comparison sites. Paired t-tests were employed to measure the potential impact of the program on reducing systolic blood pressure (SBP) and hemoglobin A1c (HbA1c) between baseline and endline, and on increasing the percentage of enrollees meeting clinical targets (SBP < 140 mmHg for hypertension; < 8% HbA1c for diabetes). We analyzed qualitative data using thematic coding.

**Results:** Across sites, 2,764 hypertension patients and 244 diabetes patients were followed through endline. Participants experienced reductions in SBP in TO (−1.9 mmHg [−3.1;−0.7]) and VC (−4,2 mmHg [−5.2;−3.1]); more hypertension patients met treatment targets in these locations (TO: +3.9 percentage-points [0.4;7.2]; VC: +10.5 percentage-points [7.81;13.2]) by endline. HbA1c decreased in TO (−0.6 [−0.9;−0.4]) and VC (−0.9 [−1.4;−0.5]), and more individuals presented HbA1c < 8% by endline (TO: +10.2 percentage-points [3.8, 16.6]; VC: +25 percentage-points [12.2, 37.8]). Qualitative data pointed to overall enthusiasm for new technologies and care routine implemented by HealthRise, but challenges regarding program implementation, integration with other levels of care, and social determinants of health persisted.

**Conclusions:** Program showed positive effects on hypertension and diabetes outcomes. Community-based health interventions can help bridge healthcare gaps, but their full impact will remain limited until multisectoral policies and actions address underlying structural and social determinants of health more effectively.

## INTRODUCTION

Non-communicable diseases (NCDs), including cardiovascular diseases (CVDs) and diabetes mellitus (DM) impose a substantial health and economic burden on populations, especially in low- and middle-income countries (LMICs) where NCD risk factors such as hypertension (HTN) and unhealthy diet are on the rise, and are a threat to sustainable development.^1^

In Brazil, a country with one of the fastest ageing populations globally,^2^ important strategic efforts^3^ aiming to tackle NCDs and its risk factors were adopted in the past decades and have successfully led to decreases in standardized NCD mortality.^4^ However, prevalence of HTN and DM are still rising. Approximately 25% and 8% of the adult population living in Brazilian state capitals are diagnosed with HTN and DM, respectively,^5^ with social aspects, such as education, income, gender, and ethnicity being associated with higher prevalence of both conditions nationwide.^6,7^ As a result, CVDs and DM account for a persistent large share of the country’s burden of disease and resulted in more than 19 million deaths in 2017.^8^

Most of the care for NCDs within the publicly funded Brazilian Unified Health System is provided through decentralized primary health care (PHC) services. Since 1994, the Family Health Strategy (FHS) focus has reorganized PHC to guarantee universal access to care and widened the scope of its action from traditional curative medical care to prevention, health promotion, and integrated practices.^9,10^ However, despite progressive expansion of the FHS – from covering 4% of the Brazilian population in 1998 to 64% in 2016 –,^9,11^ obstacles to high-quality care endure. Brazil’s PHC suffers from constrained public financing and inefficient integration with other levels of care, and many health facilities are not equipped to reliably diagnose and treat NCDs, especially those in underserved areas.^12^ As a result, sizable gaps observed along the cascade of care – diagnosing, treating, and controlling disease – emphasize the need to better reach patients, retain them in care, and promote effective interventions for disease management.^13–16^

Community-based programs, which bridge communities and health systems, have emerged as a promising approach to filling gaps in local healthcare provision.^17,18^ Past LMICs studies suggest that locally relevant community-based strategies targeting HTN and DM can contribute to improved disease management and health outcomes.^19–22^ The HealthRise program was developed to implement and evaluate pilot programs aimed to improve screening, diagnosis, management, and control of HTN and DM among underserved communities in Brazil, India, South Africa, and the US.^23,24^ In Brazil, pilot programs were implemented between 2017 and 2018 in two sites – the region of Teófilo Otoni (TO) and the city of Vitoria da Conquista (VC) – and included a range of integrated strategies aimed at reorganizing the local health system and improving FHS teams’ practices to identifying, educating, and empowering HTN and DM patients.

In this independent prospective evaluation, we used both quantitative and qualitative data to assess the potential impact of the Brazil HealthRise programs on improving clinical and health outcomes for HTN and DM patients in TO and VC. We hypothesized that participation in the Brazil HealthRise program could lead to reduced biomarker readings and increase the proportion of individuals meeting treatment targets for both conditions.

## METHODOLOGY

### Study design

HealthRise was funded by the Medtronic Foundation and the Institute for Health Metrics and Evaluation (IHME) served as the independent program evaluator. Programs were designed and implemented by local grantees. Further information on the global HealthRise initiative team structure is published elsewhere.^23,24^ we used a mixed-methods quasi-experimental design to assess the magnitude and potential effects of HealthRise programs in Brazil. We used process indicators to assess program implementation; quantitative data to measure changes in patient outcomes over time; and qualitative data to contextualize patients, providers, and stakeholders’ experiences with HealthRise and usual NCD care.

### HealthRise program

Supported by the Brazilian Ministry of Health, pilot programs were implemented in two sites: (1) TO region, a cluster of 10 municipalities located in the Mucuri Valley, in Minas Gerais (May 2017-December 2018); and (2) VC, a city located in northeastern Brazil, in Bahia state (March 2017-December 2018). Sites were selected a priori by the Medtronic Foundation and grantees due to high NCD burden, existing health service gaps, and interest of government and nongovernmental partners.

Interventions were drawn from site-specific baseline needs assessment conducted prior to program implementation and evaluation onset, which highlighted shared health demands and challenges across sites.^25^ In both locations, health infrastructure was characterized by insufficient human resources for health, no electronic medical records (EMR), medication stock-outs, and lack of equipment. Diagnosis of HTN and DM were typically delayed until after the emergence of symptoms. Once diagnosed, a majority of patients were found to initiate treatment; however, many failed to reach treatment targets. Lifestyle modifications and the time required to schedule follow-up appointments, as a result of overburdened facilities, were identified by patients as some of the major challenges to controlling their diseases.^25^

The programs’ interventions and activities covered six common components (i.e. technologies for care coordination; healthcare service organization; workforce development; screening and recruitment; disease management and health promotion; and patient empowerment and health education) and are described in more detail in Table 1.

**Table 1.** Overview of Brazil HealthRise interventions.

|  |  | Teófilo Otoni region | Vitória da Conquista |
| --- | --- | --- | --- |
| **Technologies for care coordination** | Common across sites | Equipped primary health care units with computers, notebooks, tablets, and internet; |  |
|  | Site-specific | Developed and implemented two Clinical Decision Support System for health providers' use, one for screening and one with web-based medical records which generates alerts, reminders, and an individualized therapeutic plan based on the information entered, in 34 health care units;<br>Linked health facilities to Central Telehealth Units. | Implemented Brazilian web-based medical record system (e-SUS) in 16 primary health care units;<br>Developed digital screening and job aid tools on promoting healthy lifestyles for CHWs. |
| **Healthcare service organization** | Common across sites | Purchased equipment for specialized exams (made available at secondary care level clinics) and supported teams with referral process;<br>Evaluation and optimization of human resources and patient flows for a better organization of the care for hypertension and diabetes; |  |
|  | Site-specific | Monthly web meetings with representatives of each municipality for evaluation of the processes and results of the intervention.<br>Establishment of a routine for laboratory tests, including hiring a Laboratory to visit the remote municipalities and perform the exams. | In agreement with the municipal health department, implemented one weekly shift for nurse consultations and two weekly shifts for medical consultations for patients with HTN and/or DM;<br>Hired a physician and provided evening consultations at Social Service for Industries (SESI);<br>Increased the number of specialists in cardiology, endocrinology, angiology, and ophthalmology to perform specialized exams. |
| **Workforce development** | Common across sites | Delivered in-person skill-based training on HTN and DM care management and use of new technologies to all primary care providers and community-based health worker roles;<br>Provided online courses focused on providing routine HTN and DM care, including strategies for disease management and healthy nutrition targeted to all primary care providers and community health workers. |  |
|  | Site-specific | Developed in-person training material;<br>Developed web-based courses. | N/A |
| **Screening & recruitment** | Common across sites | Performed community-based screening events. |  |
|  | Site-specific | • Implemented targeted household visits by CHWs for identification of new patients and eligible participants. | • Workplace screening events at SESI. |
| **Disease management & health promotion** | Common across sites | • Ensured follow-up with patients who missed appointments, failed to pick up medications, and/or were not meeting treatment targets;<br>• Reinforced home visits targeting patients with out of target blood pressure and glucose levels;<br>• Increased the availability of specialized exams, including ECG, echocardiogram, fundus oculi photography, and A1c point-of-care;<br>• Developed text message system to help foster adherence to treatment and regular consultations; as well as to promote healthy lifestyle habits. |  |
|  | Site-specific | • Promoted cooking workshops and physical educational sessions supported by NASF teams. | • Implemented three outdoor gyms at primary health care units. |
| **Patient empowerment & health education** | Common across sites | • Encouraged the creation, expansion, and reorganization of support groups to allow patient exchange to improve self-care and health literacy, with an emphasis on healthy nutrition and habits;<br>• Coordinated educational workshops;<br>• Developed printed educational materials to be used during consultations and home visits.<br>• Supported the creation of the municipal association for individuals with HTN and DM and assisted to engage them in the federal network of DM associations. |  |
|  | Site-specific | • N/A | • Produced educational cartoons to be aired on waiting rooms. |

### HealthRise participants

In the TO region, HealthRise activities targeted individuals aged 30-69 years old living in urban and rural areas of the selected municipalities; in the TO municipality, only those living in the same area as five selected PHCU were considered for inclusion. In VC, the target population was composed of individuals aged 30 years and above living in the urban area. Patients were enrolled in the program on an ongoing basis if (1) they had an established HTN or DM diagnosis and attended a follow-up consultation at the health facility; or (2) had no previous diagnosis, screened above threshold at any HealthRise-supported screening activity and attended a follow-up consultation at the PCHU for diagnosis confirmation. For HTN, “screened above threshold”, meant that a participant’s blood pressure (BP) exceeded diagnostic thresholds (i.e., systolic blood pressure (SBP) ≥ 140 mmHg or diastolic blood pressure (DBP) ≥ 90 mmHg). For DM, it meant a random blood glucose (RBG) measure of ≥ 140 mg/dL in VC; and a RBG reading of ≥ 200 mg/dL with at least one classical DM symptom (polyuria, polydipsia, or polyphagia) or a fasting glucose ≥ 126 mg/dL following a cardiovascular risk assessment (only individuals with a body mass index (BMI) ≥ 25; or age ≥ 45, or at least moderate cardiovascular disease risk were referred to get a fasting glucose test at the health facility) in TO. Newly diagnosed patients were those who screened above the threshold at a screening event and received a HTN or DM diagnosis after two follow-up consultations at the health facility. For those with a screening RBG reading of ≥ 200 mg/dL with at least one classical diabetes symptom or blood pressure ≥ 180 mmHg/≥ 110 mmHg one assessment was enough for diagnosis. BP and hemoglobin A1c (HbA1c) measures were used to monitor enrolled patients. Figure 1 shows information on the target population and a flowchart of participants in each of the Brazilian sites.

**Figure 1.**
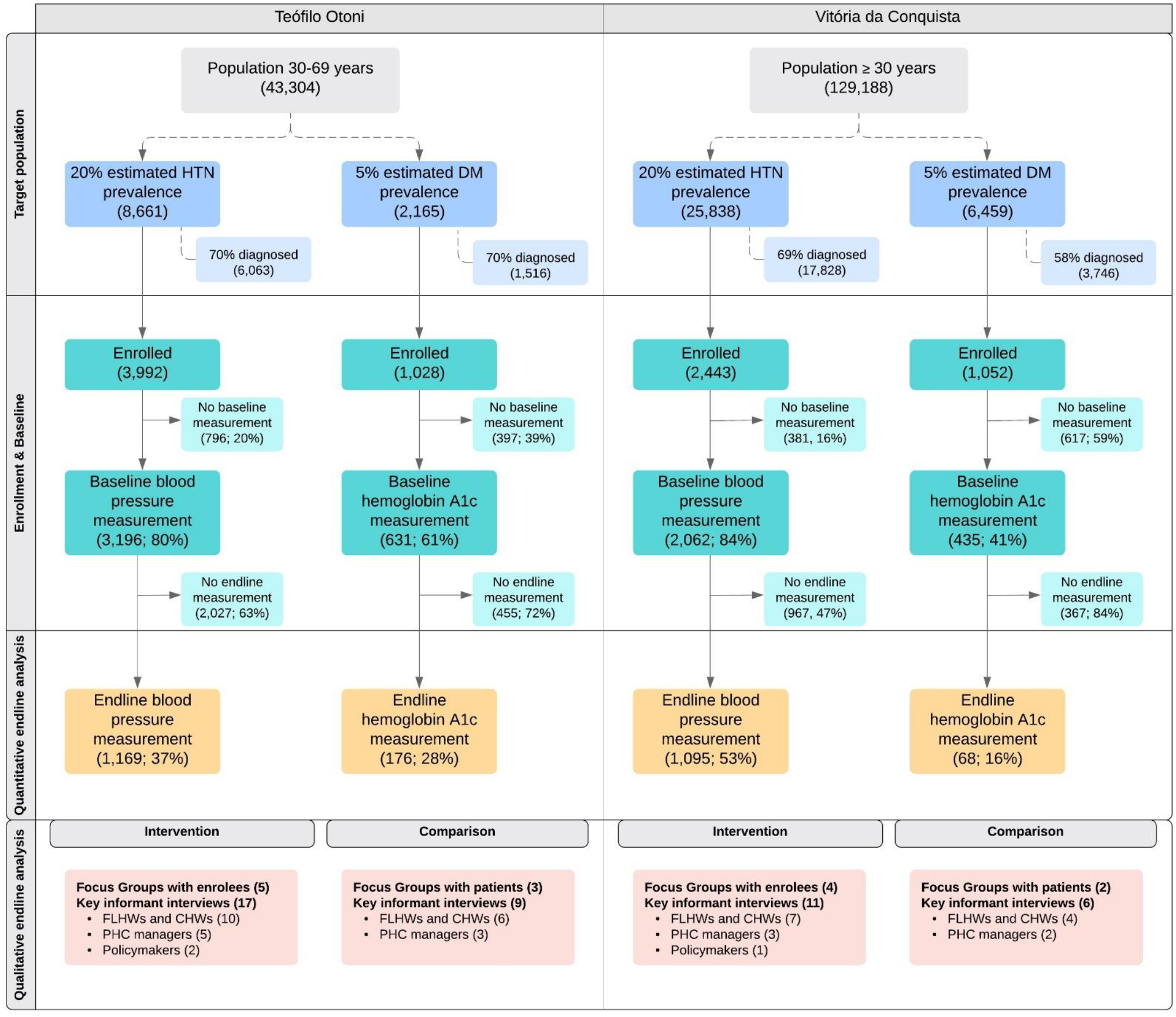
Flowchart of participants, by condition and site.

### Endline evaluation data collection

Quantitative process and patient-level data were collected from existing sources and collated by the implementation teams, who provided de-identified patient-level data for enrolled individuals over time. TO grantees used their newly implemented Clinical Decision Support System as the main source of patient-level data; while the team in VC extracted data from paper-based and electronic medical records. Data collected varied by program site because of the differing data source infrastructure.

Qualitative data were collected from both intervention and comparison areas and consisted of key informant interviews with PHC managers, clinic- and home-based providers, and policymakers, as well as focus group discussions with patients, which were facilitated by independent local data collection professionals contracted by IHME. Comparison locations – Padre Paraíso (for TO) and Poções (for VC) – were selected based on cultural, geographic, and sociodemographic proximity and the absence of any HealthRise programs. Frontline health workers (FLHWs) and CHWs were randomly selected from a list provided by each facility manager. PHC managers and policymakers were identified through the implementation teams. Patients were recruited to participate in focus group discussions by health facility staff. Figure 1 displays the number of focus group discussions and interviews conducted by site, and data collection methods are described in more detail elsewhere.^23,24^ Interviews and focus group discussions were conducted in Portuguese, audio recorded, transcribed, and translated to English. Survey instruments were designed by IHME with input from local evaluation partners and are available at http://www.healthdata.org/healthrise-evaluation/data-collection-tools.

### Endline evaluation analysis

Two outcome indicators were used to quantify the potential effects of HealthRise participation: (1) the proportion of patients meeting treatment targets (i.e., SBP < 140 mmHg and DBP < 90 mmHg for HTN; < 8% HbA1c for DM); and (2) patient biometric measures (i.e., SBP for HTN, HbA1c for DM). The endline study size was determined by the number of enrolled patients that had at least two BP or two HbA1c readings recorded (Figure 1) – one that aligned with enrollment (i.e., baseline) and any additional measure by endline. We ran paired-sample t-tests – for males, females and both sexes combined – to assess whether statistically significant changes in HTN and DM measures – percent meeting treatment targets and average biometric readings – occurred for HealthRise participants from baseline to endline. All analyses were conducted in Stata version 15 and R version 3.6.2.^26,27^

For the qualitative analysis, each transcription was read two or more times by a single researcher who assessed open-ended questionnaire responses using thematic analysis.^28^ Themes were identified at the semantic level. Data were entered into excel templates for analysis with a focus on data patterns associated with overarching research questions. Data codes were collated to generate themes by site and draw comparisons between intervention and comparison regions.

## RESULTS

Across both sites, 5,444 individuals without a previous HTN diagnosis and 9,005 individuals not previously diagnosed with DM were screened through HealthRise programs, of which, 1,497 exceeded diagnostic thresholds for elevated BP and 624 met DM referral criteria for further testing at the health facility. By the end of the programs, 493 new HTN and 84 new DM patients were diagnosed. Site-specific figures are presented in Figure 2. Females accounted for the majority of the newly diagnosed patients in TO – 52% of those diagnosed with HTN and 70% of those with DM. In VC, more males were diagnosed with HTN (53%) and more female participants received a diagnosis of DM (61%).

**Figure 2.**
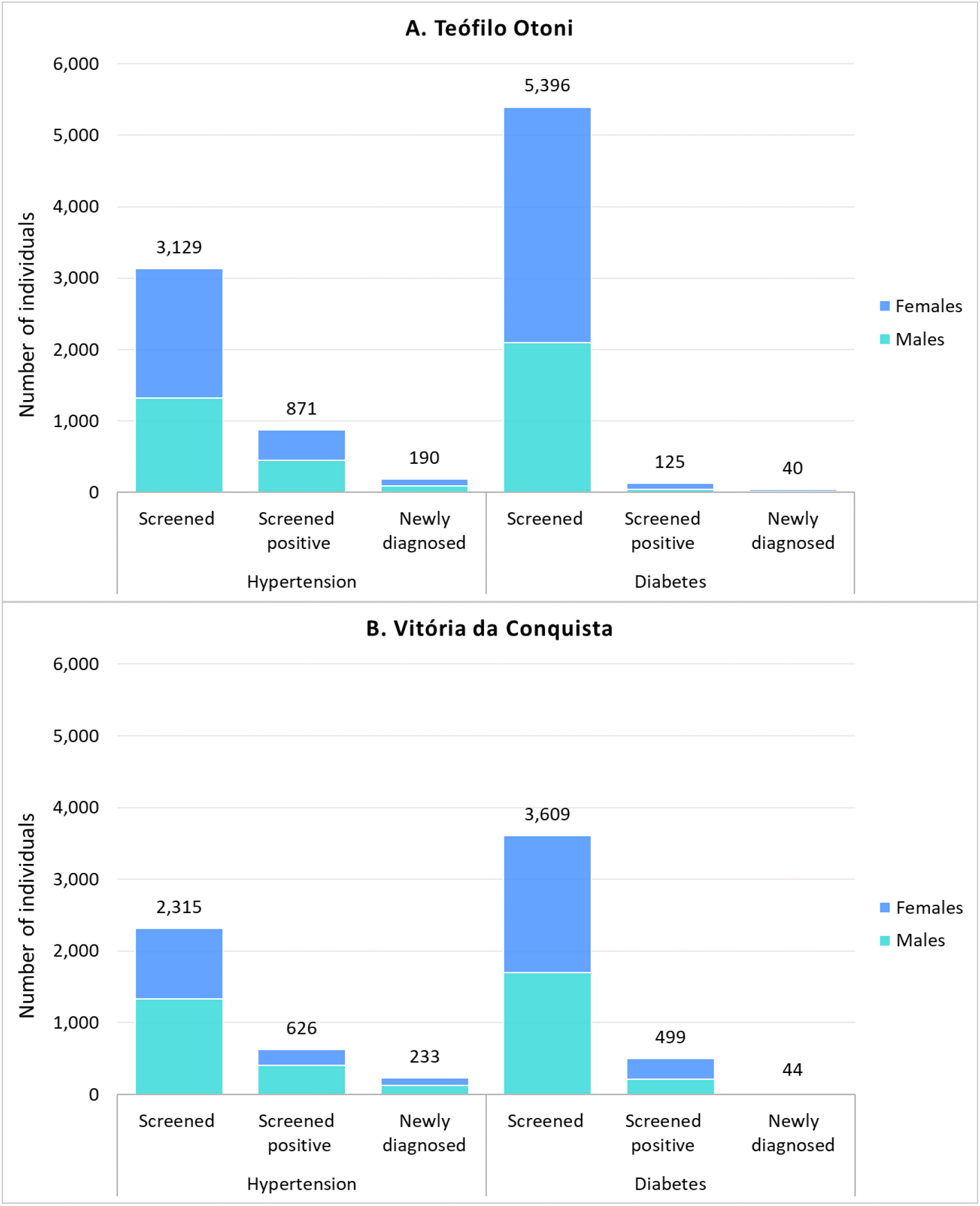
HealthRise screening and diagnosis outputs for Teófilo Otoni (A) and Vitória da Conquista (B).

A total of 3,992 individuals with HTN and 1,028 individuals with DM attended the health facility at least once and were enrolled in the program in TO; and 2,443 and 1,052 patients with HTN and DM, respectively, were enrolled in VC. Baseline BP readings were available for the majority of enrolled patients in both locations (n=3,196 [80%] in TO; 2,062 [84%] in VC), while HbA1c measurements were less frequently available (n=631 [61%] in TO; n=435 [41%] in VC). The proportion of enrollees who were achieving the recommended BP target at baseline was 48% in TO vs 35% in VC; for HbA1c the percentages were 54% and 49% for TO and VC respectively. Across sites, a considerable percentage of enrolled HTN and DM patients with a baseline biomarker reading were not followed-up and/or did not have an additional BP (63% in TO; 47% in VC) or HbA1c (72% in TO; 84% in VC) reading recorded (Figure 1).

### Changes in blood pressure and HbA1c

Figure 3 summarizes patients shifts in SBP (A) and HbA1c (B) categories between baseline and endline in TO and VC. Overall, patients trended toward progress in lowering SBP or HbA1c levels over time in both sites; however, shifts towards higher categories were also observed (Figure 3).

**Figure 3.**
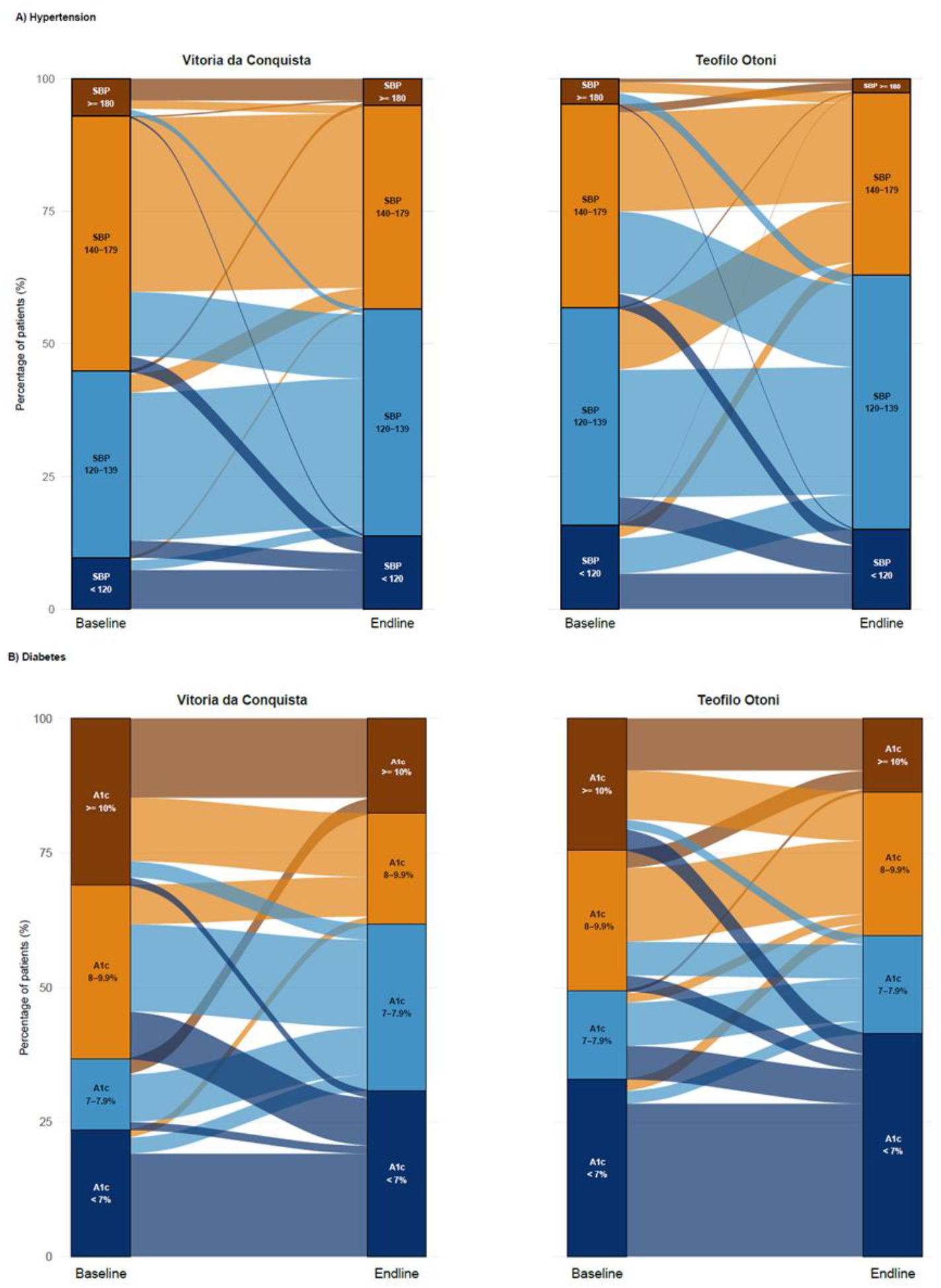
HealthRise patient shifts in biomarker ranges between baseline and endline for hypertension (A) and diabetes (B). The height of each column reflects 100% of patients at each time point (baseline and endline), while the categories within each column represents the percentage of patients in each category at baseline and endline. Patient groups are color-coded by their categorization at endline (right column per site) and flow from their categorization at baseline (left column per site).

By the end of the program, a significantly higher number of patients met HTN treatment targets at endline (52.2% [49.3;55%]) compared to baseline (48.3% [45.5;51.2%]; p < 0.05) in TO. TO HealthRise patients also recorded statistically significant reductions in SBP (average decrease of −1.9 mmHg [−3.1;−0.7]; p < 0.01), especially males. Even larger improvements occurred in VC, where a sizeable increase in the percent of HTN individuals with SBP < 140 mmHg and DBP < 90 mmHg was observed (10.5 percentage point rise [7.81;13.2; p < 0.001], and patients showed statistically significant declines for SBP (−4.2 mmHg decline [−5.2;−3.1]; p < 0.001) over the course of the intervention.

For DM, more patients met treatment targets at endline (59.6% [52.3;67.0%]) than at baseline [49.4% [42.0;56.9%]; *p* < 0.01) in TO, including 14.8% whom had A1c levels of 8% or higher at enrollment; for VC, the percentage of HealthRise DM patients meeting treatment targets also showed a statistically significantly (*p* <0.001) increase from baseline (36.8% [25.0;48.5%]) to endline (61.8% [49.9;73.6%]). Statistically significant increases were seen among female participants only in both locations. Improvements were also observed in terms of reducing HbA1c since enrollment, with an average decrease of −0.6 ([−0.87;−0.36]; *p* < 0.001) in TO and of −0.9 ([−1.4;−0.5]; *p* <0.001) in VC. No significant changes in mean HbA1c levels were seen among male participants in TO.

### Qualitative findings

Key themes emerged from the qualitative data collected at the intervention sites (Table 2). First, providers described the program as reorganizing patient flows and health unit routines, resulting in better and more structured care delivery. Providers also spoke positively about new training opportunities, intensified group activities for patients, and an increased availability of select specialized exams. In TO, specifically, the implementation of the clinical support system resulted in an improved sense of confidence among frontline health workers according to providers, and was highlighted as one of the main features of the program. Second, while patients demonstrated limited understanding of HealthRise, they were able to recognize some of the main program aspects, such as screening health fairs and some educational activities. The use of tablets by CHWs during home visits and computers by FLHWs during consultations was also perceived positively by patients. Individuals from TO more frequently reported enhanced access to specialized exams after the implementation of HealthRise.

**Table 2.** Summary of key themes, components, and quotes from qualitative data synthesized across Brazil HealthRise sites.

| HealthRise thematic area component and contexts | Sample thematic quotes |
| --- | --- |
| **Theme 1: Program strengths** |  |
| Improved provider experience from reorganized patient flows, new training opportunities, and increased availability of exams;<br>Introduction of tablets to aid in patient record-keeping and care decisions in Teófilo Otoni;<br>Implementation of Electronic Medical Records in Vitória da Conquista. | *"HealthRise has brought some changes to our work process. It has changed some of the things that were not working before."* – FLHW<br>*"I think the EMR resulted in a better way of communicating about the patient...any professional can now access the information stored in there."* – FLHW<br>*"With the Decision Support System I feel more confident now, and I know exactly what I need to do."* – FLHW<br>*"In my opinion, one of the main goals of the project was to bring back to the system those patients who didn't use to care about their diabetes or hypertension."* – CHW |
| **Theme 2: Patients experiences with HealthRise** |  |
| Screening events and educational activities identified as HealthRise-specific activities;<br>Increased access to specialized exams;<br>Positive view of new technologies for care coordination (i.e. tablets and computers). | *"They got a device that you take home with you to monitor your blood pressure."* – Patient<br>*"The health fair was really good. There were some educational activities in there."* – Patient<br>*"It was because of this project that I found out about my condition."* – Patient<br>*"I was even able to get my eyes examined!"* – Patient<br>*"Now I feel more confident. We need to go to the health unit frequently, to get some tests frequently, you need to collect blood more frequently as well."* – Patient<br>*"They didn't use computers before and now they use it. And there is everything in there...the frequency we go to the health unit, the medication we take..."* – Patient |
| **Theme 3: Remaining challenges** |  |
| Impact of social determinants of health on NCD risk, onset, and treatment;<br>Barriers to disease management from local food culture and health system constraints;<br>Moderate-low knowledge about NCDs and low adherence to therapeutic plan | *"I really like eating rice, but we can't. But I eat it anyway."* – Patient<br>*"The main challenge is to bring the patient to the health unit."* – FLHW<br>*"Not always we can refer a patient to the nutritionist or to the physiotherapist, there are too many patients to such a reduced team."* – FLHW<br>*"The main barrier is to not have a doctor here every day. There is no pharmacist."* – Facility manager |
| **Theme 4: Recommendations for improvement** |  |
| • Additional training opportunities;<br>• Strengthened care coordination with a focus on multidisciplinary approaches;<br>• To overcome health systems obstacles to high quality care. | *“The trainings were good, but they need to be permanent...” – FLHW*<br>*“A more intense multidisciplinary approach...I missed that a lot. A psychologist, a nutritionist...so that we could discuss the cases together.” – FLHW* |

The third theme pertained to remaining challenges for disease management. Social determinants of health, including poverty, low levels of health education, and limited access to affordable and nutritious food were consistently reported across sites. Patients, providers, and policymakers also highlighted challenges related to core health system functions, particularly adequate medication supplies, sufficient staffing, reliable referral processes, and budget limitations within the public health system.

Lastly, in both sites, FLHWs and CHWs requested additional and more periodic in-person training opportunities. Specifically, CHWs frequently expressed interest in in-depth technical trainings that would allow them to measure blood glucose and BP during home visits. Interviewees also expressed the desire for a strengthened care coordination, with health professionals other than doctors and nurses to be more closely involved in the project to support a truly multidisciplinary approach. Finally, guaranteeing medication availability and increasing access to specialized exams at the local level, without the need to refer patients to a central unit in a different municipality, would be key improvements to the program and the local health systems.

Patient focus group discussions and staff interviews at comparison facilities echoed many of the same themes as those from intervention areas. However, while complaints regarding the availability of medication and specialized exams also arose in intervention facilities, these complaints were clearly more escalated in comparison areas. Although facilitating the referral process to specialists was a core component of HealthRise in both sites, this seemed to be a major barrier in comparison facilities, according to patients, health providers, and PHC managers. Additionally, FLHWs in comparison areas agreed that HTN and DM were not their main focus given other local competing health priorities (i.e. maternal and child health). Patient adherence to standard HTN and DM programs is low, according to health staff. Finally, providers in comparison facilities more frequently indicated the need for additional training opportunities.

## Discussion

The HealthRise program focused on overcoming barriers to effective care and improving clinical and health outcomes for HTN and DM among underserved populations through community-based interventions. Findings from this evaluation reveal that enrolled patients followed through endline saw statistically significant reduction in blood pressure, and the proportion of patients meeting HTN treatment targets increased in both sites, despite the short period of program implementation. Similarly, HealthRise patients’ HbA1c levels significantly decreased and patients experienced improvements in DM management since baseline in both TO and VC. Observable effects varied across genders and statistically significant positive trends were more frequently seen among women compared to men. Results from the qualitative assessment pointed to the value of integrating interventions covering health promotion, prevention, and treatment, and of strategies focused on healthcare service organization; nonetheless, gaps related to core health system functions as well as socioeconomic and cultural barriers to care provision and disease management were identified by providers, managers, and patients from both intervention and comparison facilities.

Unlike most primary care interventions,^18^ HealthRise covered multiple domains, from organizing and equipping healthcare facilities and teams to community-level actions and policymaking efforts, which may have contributed to the observable effects of HealthRise across Brazil sites. A review of comprehensive chronic care models indicated that the majority of them were effective in improving healthcare practices and health outcomes within primary healthcare settings.^29^ In Brazil, despite the lack of previous overarching multicomponent efforts added to standard care, a number of programs that have implemented at least one of the interventions covered by HealthRise were effective in improving blood pressure and blood glucose measures, but at smaller levels.^30–32^ Further, the country’s longstanding investments in guaranteeing universal healthcare coverage, strengthening primary care and establishing one of the world’s largest CHW networks^33,34^ may have contributed to the notable progress and may explain the lack of statistically significant results in other HealthRise implementation sites.^23^

Amid such promising findings, however, several operational challenges arose over the course of the Brazilian programs and can inform future community-based interventions, especially those in underserved communities. HealthRise programs screened thousands of individuals, yet relatively few new diagnoses occurred through this process. Low yields from population-based screening activities are not uncommon.^35,36^ Given the estimated size of undiagnosed patients in each implementation site, large drop-off rates between screening above thresholds and then receiving diagnostic confirmation via multiple clinic appointments may have contributed, at least partially, to the low numbers, a challenge also identified elsewhere.^35^ These findings support guidelines recommending more selective screening of high-risk groups to improve cost-effectiveness.^37^

As in other community-based interventions in resource-constrained settings,^38,39^ low follow-up rates were observed in Brazil. Although the short time of implementation may have limited the amount of HealthRise patients with multiple biomarker readings, persistent health systems barriers raised by providers and managers in qualitative analyses, such as overburdened facilities, limited staff, and lack of diagnostic tools, likely contributed to these findings. As such, innovative HTN and DM programs need to develop better strategies to reach patients, increase follow-up opportunities, reduce delays in medical appointments, and retain enrolled patients. Strengthening CHWs technical abilities and promoting task shifts can potentially alleviate some of these barriers and better support patients’ needs. However, further work is needed to understand the capacity of CHW to incorporate these other demands while dealing with both persistent and emergent – such as COVID-19 – competing health priorities.

In both sites, males represented a smaller share of enrolled and followed-up patients, indicating that some access barriers – especially facility working hours – might disproportionally affect males. Although the smaller sample size may have impacted the results for males, gender-specific health barriers and differences regarding healthcare-seeking behavior and health literacy are well established in the literature.^40,41^ A combination of these aspects may have led to insufficient exposure to the program’s interventions and compromised management of chronic conditions among male participants indicating that gender may need to be considered when designing similar interventions.

Lastly, across sites, heterogeneous patterns in patient biometrics shifts since baseline underscore the challenges of case management of HTN and DM, especially in environments with the aforementioned health system fragilities and where social and cultural aspects heavily interfere with access to care and adoption of the therapeutic plan, exacerbating health disparities. Interviewees have pointed to persistent obstacles to accessing multidisciplinary care, unreliably stocked medication, and lack of integrated care with specialized levels and providers, which may have resulted in some hard-to-treat patients remaining in suboptimal treatment and thus not achieving better disease management results. Identifying and prioritizing mechanisms for mitigating or alleviating structural inequalities is mandatory so programs can reach their full potential. Given that CHW are reported to be trusted and effective in reaching families and providing ongoing assistance while also having a unique understanding of the experience, language, culture, and socioeconomic reality of the communities that they serve, they are vital in the path towards health equity.

### Limitations

Our study’s findings should be interpreted in light of its limitations. First, since quantitative comparison patient data were not collected, we cannot ascribe these patient-level improvements to HealthRise program participation. Second, only patients with two or more biomarker readings by endline were included in the analysis; these patients were considered to have been regularly followed up and may not be representative of the potential target population for HealthRise interventions (i.e. bias toward healthier or sicker patients; or towards females). There is also a chance that patients ended up excluded from the analysis due to poor record keeping. Third, our relatively small sample size, especially of DM patients, alongside with data availability limitations precluded additional sub-group analysis as well as investigations regarding visit frequency, intensity of intervention exposure, and medication use. Fourth, comprehensive information on intervention reach and fidelity (i.e., the degree to which interventions were implemented per protocol) were not available across sites and thus could not be included in the present study. Finally, given the open enrollment, patient duration in the program varied and, for some, may not have been long enough to detect significant effects.

## Conclusions

Our findings demonstrate the potential for community based interventions to improve HTN and DM outcomes; but also underscore how their reach and effectiveness can be hindered by broader health system, infrastructure, and policy constraints. Longer-term evaluations of community-based programs and continued work to understand which interventions may work best given local contexts and healthcare gaps are needed. Irrespective of their increasingly vital role for underserved populations, community-based programs can be hindered by the lack of underlying infrastructure and resources and absence of more macro-level socioeconomic policies. It is imperative that models of care explicitly tackle local contextual challenges and engage well beyond the formal health system.

## Ethics and consent

The overall study was approved by the Institutional Review Board of the Human Subjects Division at the University of Washington. (Application Number: 51476) Institutional Review Board approval was also obtained for both quantitative and qualitative data collection from the Universidade Federal dos Vales do Jequitinhonha e Mucuri (CAAE:65808517.9.0000.5108 and CAAE:00865518.9.00005108) and the Universidade Federal da Bahia, Instituto Multidisciplinar em Saúde-Campus Anísio Teixeira (CAAE:62259116.0.0000.5556 and CAAE:99216918.5.0000.5556). All personal identifiers were removed prior to the data being sent to IHME for analysis. Individuals participating in the study in TO provided informed consent. Quantitative data from participants in VC were extracted from patient charts and no informed consent was needed. All those participating in qualitative data collection activities provided informed consent.

## Data Availability

The datasets used and/or analyzed during the current study are available from the corresponding author on reasonable request.

## Notes

### Competing Interest Statement

PB and JD are employees of the Medtronic Foundation. MTUB is a former Medtronic Foundation advisor. WWA, VMB, MALB, CCRC, MLC, PWE, CNK, MMOL, JAL, JXM, MSM, DSM, SM, JAQO, MGO, VSOAP, ALPR, DR, KOS, and DAS are recipients of HealthRise grants from the Medtronic Foundation to implement HealthRise interventions. LSF, SW, JNC, NF, KPH, CRM, MN, BKP, BT, and EG are recipients of funding from grants from the Medtronic Foundation to evaluate HealthRise interventions.

### Funding Statement

Funding for the HealthRise project came from the Medtronic Foundation. The corresponding author had full access to all the data in the study and had final responsibility for the decision to submit for publication. This study was financed in part by the Coordenacao de Aperfeicoamento de Pessoal de Nivel Superior - Brazil (CAPES) - Finance Code 001. JAQO and ALPR received support from the Instituto de Avaliacao de Tecnologia em Saude (IATS) and the Conselho Nacional de Desenvolvimento Cientifico e Tecnologico (CNPq), Brazil. ALPR was supported in part by Fundacao de Amparo a Pesquisa do Estado de Minas Gerais (FAPEMIG). MGO received funding from the Bahia State Research Support Foundation, the Federal University of Bahia, and the Municipal Health Secretariat of Vitoria da Conquista.
Funders had no role in the study design, data collection, analysis, interpretation of data, or writing of the first draft of this manuscript.

### Author Declarations

The overall study was approved by the Institutional Review Board of the Human Subjects Division at the University of Washington. (Application Number: 51476) Institutional Review Board approval was also obtained for both quantitative and qualitative data collection from the Universidade Federal dos Vales do Jequitinhonha e Mucuri (CAAE:65808517.9.0000.5108 and CAAE:00865518.9.00005108) and the Universidade Federal da Bahia, Instituto Multidisciplinar em Saude-Campus Anisio Teixeira (CAAE:62259116.0.0000.5556 and CAAE:99216918.5.0000.5556). All personal identifiers were removed prior to the data being sent to IHME for analysis. Individuals participating in the study in Teofilo Otoni provided informed consent. Quantitative data from participants in Vitoria da Conquista were extracted from patient charts and no informed consent was required. All those participating in qualitative data collection activities provided informed consent.

